# Rapid progression to death after hepatocellular carcinoma diagnosis particularly among persons with advanced HIV disease in Kampala, Uganda

**DOI:** 10.1101/2022.06.24.22276850

**Authors:** Sara K. Nsibirwa, Jim Aizire, David L. Thomas, Ponsiano Ocama, Gregory D. Kirk

**Affiliations:** HIV and HCC in Uganda (H^2^U) Consortium, Kampala, Uganda; Infectious Diseases Institute (IDI), Kampala, Uganda; Johns Hopkins University, Baltimore, MD, USA; Makerere University College of Health Sciences, Kampala, Uganda; Uganda Cancer Institute, Kampala, Uganda

**Keywords:** hepatocellular carcinoma (HCC), mortality, Human Immunodeficiency Virus, sub-Saharan Africa

## Abstract

**Background:** HIV infection is associated with more rapid progression of some comorbidities. This study assessed the impact of HIV-infection where the presentation and outcome of HCC was different in persons living with HIV (PLWH) compared to those without HIV infection.

**Methods:** HCC patients attending the Mulago National Referral Hospital in Uganda were enrolled into a natural history study of HCC between March 2015 and February 2019. Standardized methods were used to collect clinical, ultrasound and laboratory data at enrolment. HCC cases were confirmed based on a combination of clinical, ultrasound, tumor marker and pathology data. Follow-up contact was made at one, three, six, and twelve months post-enrolment to determine vital status. Symptoms and signs at diagnosis and subsequent survival were compared by HIV status. Kaplan Meier curves were used to assess HCC survival.

**Results:** Of 441 persons with HCC, 383 (87.0%) died within 12 months following HCC diagnosis. The median (IQR) survival was 42 (20, 106) days. The most commonly reported symptom clusters were pain (80%), gastrointestinal symptoms (28%) and anorexia / cachexia (10%), and no differences were detected in these presenting symptoms (nor most other initial findings) in the 79 (18%) PLWH compared to those without HIV. After adjusting for baseline demographic and clinical characteristics, HIV infection was associated with increased mortality but only among those with severe HIV-associated immunosuppression (CD4 count <200 cells per cubic milliliter), aHR (95% C) = 2.12 (1.23-3.53), p=0.004, and not among PLWH with ≥200 CD4 cells per cubic milliliter, aHR (95% C) = 1.15 (0.82-1.60), p=0.417.

**Conclusion:** Among relatively young Ugandans, HCC is a devastating disease with rapid mortality that is especially rapid among PLWH. HIV was associated with slightly higher mortality, notably among PLWH with lower CD4 cell counts. As a substantial majority of PLWH diagnosed with HCC were engaged in HIV care, further investigation should determine the effectiveness of incorporating screening and early identification of HCC among high-risk individuals into existing HIV care programs. Concurrent with growing access to curative localized treatment for HCC in sub-Saharan Africa, leveraging HIV care infrastructure affords opportunities for earlier HCC intervention.

## Introduction

Hepatocellular carcinoma (HCC) is the sixth most commonly diagnosed cancer and the third leading cause of cancer death worldwide.^1^ It is one of the least curable malignancies with the number of new cases reported (905,677) approximating the number of deaths (830,180) worldwide in 2020.^1,2^ HCC has the amongst the shortest survival time of any cancer, ^3^ with an even poorer prognosis among patients in lower-income countries which disproportionately bear a higher burden of primary risk factors and of HCC.^4^ With the advent of effective combination antiretroviral therapy (cART), people living with HIV infection (PLWH) have had dramatically improved survival.^5,6^ While the rates of many AIDS-defining cancers have declined, non-AIDS defining cancers including HCC are increasing in incidence.^7,8^ Previous studies looking at incidence of HCC during the cART-era have shown an increase in HCC incidence in HIV-infected persons, compared with the general population.^9,10^ Trends in North America have demonstrated increased HCC incidence rates among PLWH coinciding with increased uptake and efficacy of ART.^11^ These data raise the question of the impact that this transition may have in sub-Saharan African which accounts for more than 70% of the global HIV burden, where cART is being widely rolled out and where the median survival for PLWH is increasing.^12^ Further, compared to more developed regions, sub-Saharan Africa has higher underlying HCC incidence rates and greater burden of HCC risk factors (e.g., endemic hepatitis B virus (HBV) infection, broad aflatoxin exposure) which may substantially influence the burden of HCC among PLWH in Africa.

In more developed regions, PLWH with cancer experienced higher cancer-specific mortality than HIV-uninfected patients, independent of cancer stage or receipt of cancer treatment.^13^ General estimates of cancer survival in Uganda have shown a poor prognosis for patients with a 2.3% 5-year survival rate.^14^ Earlier studies conducted on a cohort of Ugandan HCC patients showed a median survival of 1 month among untreated patients.^15^ However, these data are over 30 years old and predate the current situation of substantial HIV prevalence of 6.5%^16^ among adults. As a result, there are a paucity of data in Uganda on the impact of HIV infection on the clinical presentation and subsequent survival of HCC cases.

The objective of this study was therefore to analyze data on the clinical presentation at HCC diagnosis and characterize survival in a contemporary cohort of patients diagnosed with HCC in Uganda, emphasizing any differences attributable to HIV infection.

## Methods

### Study population

As part of a hospital-based case-control study, HCC patients were prospectively recruited from the gastrointestinal and oncology wards of Mulago National Referral Hospital in Kampala, Uganda between March 2015 and February 2019. Potential participants were deemed eligible if they were 12 years of age or older and spoke and comprehended English or Luganda, the languages in which consenting materials were administered. All adult participants provided written informed consent; assent and parental/guardian consent were obtained prior to enrolling 12-17-year-old individuals. The study was approved by the Makerere University School of Medicine Research and Ethics committee, Uganda National Council for Science and Technology and the Johns Hopkins University Institutional Review Board.

### Study procedures

All enrolled participants underwent standardized interview, clinical, and ultrasound examination. Questionnaires were administered by trained staff and collected data on socio-demographic, behavioral and clinical factors, and the duration and type of presenting symptoms. Patients self-reported their initial symptom(s) and indicated their duration. Further, interviewers assessed symptoms commonly occurring in HCC at time of enrollment utilizing a standardized questionnaire previously used in HCC studies from Africa.^17,18^ Clinical examination assessed physical signs reflective of HCC or advanced liver disease including jaundice, wasting, hepatosplenomegaly, abdominal tenderness, ascites, collateral veins, hepatic bruit and asterixis. These data were collected by study physicians who underwent standardized training using uniform data collection instruments.

Laboratory testing was performed at a single laboratory (Makerere University-Johns Hopkins University Core Laboratory; Kampala, Uganda) for hepatitis B surface antigen (HBsAg), anti-hepatitis B core antibodies (anti-HBc), anti-hepatitis B surface antibodies (anti-HBS), anti-hepatitis C antibodies (anti-HCV), HIV antibody, alpha-fetoprotein (AFP), alanine aminotransferase (ALT), aspartate aminotransferase (AST), total bilirubin, international normalized ratio (INR), and a complete blood cell count. Urine was tested for circulating cathodic antigen (CCA) for diagnosis of active *Schistosomiasis* infection.

Ultrasound was performed by trained radiographers/radiologists with data collected on a standardized ultrasound form, as previously described.^17,19^ For this analysis, characterization of tumor number and size were incorporated.

#### HCC case definition

Participants with a clinical presentation compatible with HCC were confirmed by a standardized ultrasound examination showing space-occupying lesions with either an AFP level ≥100 ng/ml or liver biopsy documenting histologically-confirmed HCC. Study staff facilitated the completion of diagnostic assessments and almost uniformly, the initial diagnostic confirmation of HCC was concurrent with study enrollment. No HCC cases were identified as part of routine HCC surveillance but were evaluated due to clinical symptoms.

#### Survival follow-up

Contact information of both the patient and of secondary contacts was obtained at enrollment and used for participant tracing, primarily by telephone, to ascertain vital status at 1,3, 6 and 12 months after study enrolment. For individuals that were not reachable following three attempted telephone contacts, field tracing was carried out using address information. Using these follow-up procedures, vital status was verifiable for >95% of HCC cases, resulting in 419 HCC cases analyzable at three months or beyond for survival outcomes. Additional information obtained included whether the patient was hospitalized or not, date of death and reported cause of death obtained from death certificates if available to informant. Vital status and cause of death information was also determined by reviewing hospital records for those who were hospitalized.

### Statistical analysis

Pearson chi-squared (*χ*^2^) and Wilcoxon rank-sum (Mann-Whitney) tests were used to compare baseline categorical and continuous variables (e.g., demographics, symptoms, signs), respectively, stratified by HIV status. The primary outcome measure in this analysis was mortality risk following HCC diagnosis (study enrolment was used as a proxy). Kaplan-Meier methods were used to compare survival curves by key factors of HIV status and HIV-associated markers. Cox-proportional hazards regression was used to estimate the crude and adjusted hazard ratios (HR) and corresponding 95% confidence intervals (95% CI) for mortality risk. The multivariable model assessed the association between HIV status and mortality risk controlling for potential confounders defined *a priori* including age, sex, education, history of alcohol use, history of cigarette smoking, HBsAg, anti-HCV, and urine *Schistosomiasis* CCA. All *p*-values were based on a two-sided hypothesis test with a type-1 error (α = 0.05). Stata 13.0 (College Station, TX) was used for statistical analysis.

## Results

Characteristics of 441 HCC cases are shown in Table 1, overall and stratified by HIV status (N= 79, 18.0% were HIV-infected). Two-thirds of HCC cases were male, and the median age was 42 years (inter-quartile range, [IQR] 31-55 years). Almost half reported a history of alcohol intake, while 82 (19.0%) reported history of cigarette smoking. Co-infections were common: 192 (43.9%) were HBsAg positive, 30 (6.9%) were anti-HCV positive, and 214 (52.8%) were urine CCA positive for active *Schistosomiasis* infection. Overall, 219 (50.3%) of cases had elevated AFP concentrations ≥ 100 IU/ml), while 344 (78.2%) had serum AST levels above 45 U/l, 244 (55.4%) had serum ALT levels above 40 U/l, and INR was raised above the upper limit of normal (1.15) among 351 (82.2%) of cases, and three-quarters of cases had multifocal disease by ultrasound.

**Table 1:** Characteristics of HCC cases at diagnosis, stratified by HIV status.

| Characteristics | Overall*<br>N=441 | HIV positive<br>n=79 (18.0%) | HIV negative<br>n=361 (82.0%) | p-value |
| --- | --- | --- | --- | --- |
| **Socio-Demographic** |  |  |  |  |
| Age (years), median (IQR) | 42 (31-55) | 45 (37-52) | 42(30-56) | 0.463<sup>a</sup> |
| Age groups, n (%) |  |  |  | **0.001<sup>b</sup>** |
| 30 years or younger | 104 (23.6) | 9 (11.4) | 95 (26.3) |  |
| 31-50 years | 187 (42.5) | 48 (60.8) | 139 (39.0) |  |
| Above 50 years | 149 (33.9) | 22 (27.9) | 127 (35.2) |  |
| Males, n (%) | 295 (67.1) | 51 (64.6) | 244 (67.6) | 0.600<sup>b</sup> |
| Education attained |  |  |  | **0.034<sup>b</sup>** |
| Primary or less | 233 (53.0) | 52 (65.8) | 181 (50.1) |  |
| Secondary | 139 (31.6) | 20 (25.3) | 119 (33.0) |  |
| Tertiary | 68 (15.4) | 7 (8.9) | 61 (16.9) |  |
| Alcohol consumption, n (%) | 194 (44.1) | 48 (60.8) | 146 (40.4) | **0.001<sup>b</sup>** |
| Cigarette smoking, n (%) | 82 (19.0) | 22 (28.0) | 60 (17.0) | **0.025<sup>b</sup>** |
| **Clinical** |  |  |  |  |
| Physical activity, Karnofsky score, < 50, n (%) | 44 (10.1) | 10 (12.7) | 34(9.5) | 0.39<sup>b</sup> |
| Hand grip strength, > 45 pounds, n (%) | 188 (47.0) | 31 (41.3) | 157 (48.2) | 0.310<sup>b</sup> |
| **Laboratory** |  |  |  |  |
| HBsAg positive, n (%) | 192 (43.9) | 35 (44.9) | 157 (43.6) | 0.900<sup>b</sup> |
| Anti-HCV positive, n (%) | 30 (6.9) | 2 (2.5) | 28 (7.8) | 0.137<sup>b</sup> |
| Active *Schistosomiasis* (Urine CCA) positive, n (%) | 214 (52.8) | 37 (50.0) | 177 (53.5) | 0.610<sup>b</sup> |
| AST (U/l), >45, n (%) | 344 (78.2) | 69 (87.3) | 275 (76.2) | **0.035<sup>b</sup>** |
| ALT (U/l), >40, n (%) | 244 (55.5) | 48 (60.8) | 196 (54.3) | 0.319<sup>b</sup> |
| INR >1.15, n (%) | 351 (82.2) | 66 (86.8) | 285 (81.2) | 0.321<sup>b</sup> |
| AFP (IU/mL), ≥100, n (%) | 218 (50.2) | 39 (50.0) | 179 (50.3) | 1.000<sup>b</sup> |
| AFP (IU/mL), ≥400, n (%) | 199 (45.9) | 33 (42.3) | 166 (46.6) | 0.532<sup>b</sup> |
| HIV load (copies/ml) <sup>c</sup> |  |  |  |  |
| <200 | -- | 51 (64.6%) | -- |  |
| ≥200 | -- | 28 (35.4%) | -- |  |
| CD4 cell count (cells/mm3) <sup>d</sup> |  |  |  |  |
| <200 | -- | 21 (26.6%) | -- |  |
| ≥200 | -- | 58 (73.4%) | -- |  |
| **Ultrasound imaging** |  |  |  |  |
| Multifocal disease, n (%) | 316 (73.6%) | 61 (77.2%) | 255 (72.9%) | 0.730<sup>b</sup> |

Key: AFP, alpha-fetoprotein; ALT, alanine aminotransferase; anti-HCV, hepatitis C virus antibody; AST, aspartate aminotransferase; CCA, circulating cathodic antigen, a urine-based, point-of-care test for active *Schistosomiasis* diagnosis; HBsAg, hepatitis B surface antigen; INR, international normalized ratio, a measure of how long it takes blood to form a clot; IQR, interquartile range; <sup>a</sup> p-value from Wilcoxon rank-sum test; <sup>b</sup> p-value from Fisher's exact test. Highlighted p-values are statistically significant. <sup>c</sup> HIV viral suppression defined as less than 200 copies of HIV per millilitre of blood. <sup>d</sup> HIV severe disease defined as less than 200 CD4 cells per cubic millilitre). \*Missing data: AFP (n=6); anti-HCV (n=2); AST (n=1); grip strength (n=39 (8.8%)); HBsAg (n=2); HIV (n=1); INR (n=13); Multifocal disease (n=11)

HIV-associated HCC cases were a median of three years older (45 vs. 42 years, P=0.62) although their age range was notably more restricted with almost two-thirds between 31-50 years of age compared to non-HIV HCC cases with an equal proportion in the extremes of age (P= 0.001). HIV-associated HCC cases were less educated, reported more prior alcohol use and cigarette smoking, and had higher AST levels at diagnosis. Among the HIV-associated cases, 69 (87.3%) had been diagnosed with HIV prior to study enrollment. The median (IQR) CD4 count was 308 (5-1244) cells/ml^3^ among all HIV-associated cases, 316 (25-1244) among those with known HIV infection prior to HCC diagnosis, and was 262 (5-1005) among those not receiving cART. Of the 69 cases with known HIV infection, 58 (84.1%) were on some form of cART and 32 (58.2%) of these reported consistent use of cART in the month prior to study enrolment. Overall, 51 (64.6%) had an HIV viral load less than 200 copies /ml while among those receiving ART 45 (77.6%) were virally suppressed at this cutoff.

### Clinical presentation

At enrollment, most HCC cases reported multiple symptoms (Figure 1, Panel A). Pain (95%) and weight loss (93%) were almost uniformly reported, followed by abdominal fullness and anorexia in over 80% of participants. As shown in Figure 1, the frequency and patterns of reported symptoms at study enrollment were similar for both HIV-infected and HIV-uninfected cases, although more reports of nausea were reported by the HIV-uninfected cases (p=0.05). Similarly, in terms of clinical signs noted on standardized clinical examination, HIV-associated HCC cases were similar to non-HIV HCC except that flapping was exhibited more frequently among the HIV-infected cases (p=0.004), (Figure 1, Panel B).

**Figure 1:**
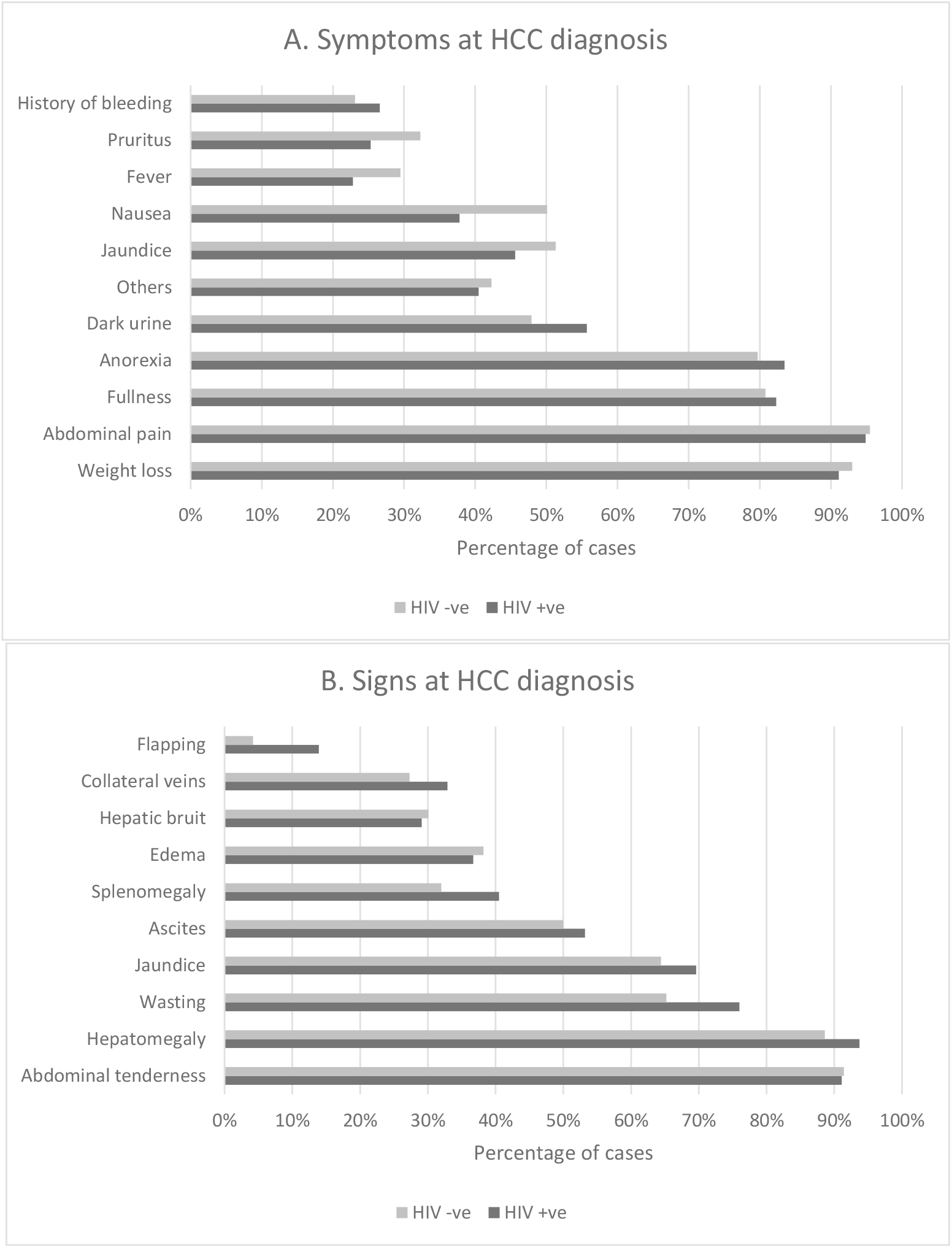
Symptoms (A) and signs (B) of liver diseases at hepatocullar carcinoma diagnosis stratified by HIV infection status. Figure 1 A - Symptoms reported by patients at HCC diagnosis, stratified by HIV infection. Figure 1 B-Clinical signs exhibited by patients on examination at time of diagnosis; stratified by HIV infection.Patients were examined for clinical signs of liver disease and HCC.

### Survival among HCC cases

Through 12 months follow-up post-HCC diagnosis with an accumulated 1,209 person-months at risk, vital status was verifiable for all but 22 (5.0%) HCC cases; 4 (5.0%) of which were HIV-infected. There were no significant differences between the participants whose vital status was ascertained at 12 months post HCC diagnosis versus those who were lost to follow-up regarding the distribution of baseline characteristics.

Kaplan-Meier risk curves overall and by HIV status and severity of immunosuppression at diagnosis are depicted in Figure 2. The overall median observed survival time was only 42 days (IQR, 20, 106) after HCC diagnosis. In comparison of HIV-associated HCC to non-HIV HCC, the median survival was 29 days (IQR, 16, 85) and 45 days (IQR, 20, 112), respectively. Overall, cumulative risk of death by 1, 3, 6 and 12 months was 42.6%, 72.7%, 84.7% and 90.7%, respectively; which was higher in the HIV versus non-HIV groups at earlier timepoints: 137 54.4% vs. 44.6% at 1 month (P 0.13), 75.9% vs. 70.1% at 3 months (P 0.47), and 84.8% vs. 81.7% at 6 months (P 0.77), respectively.

**Figure 2:**
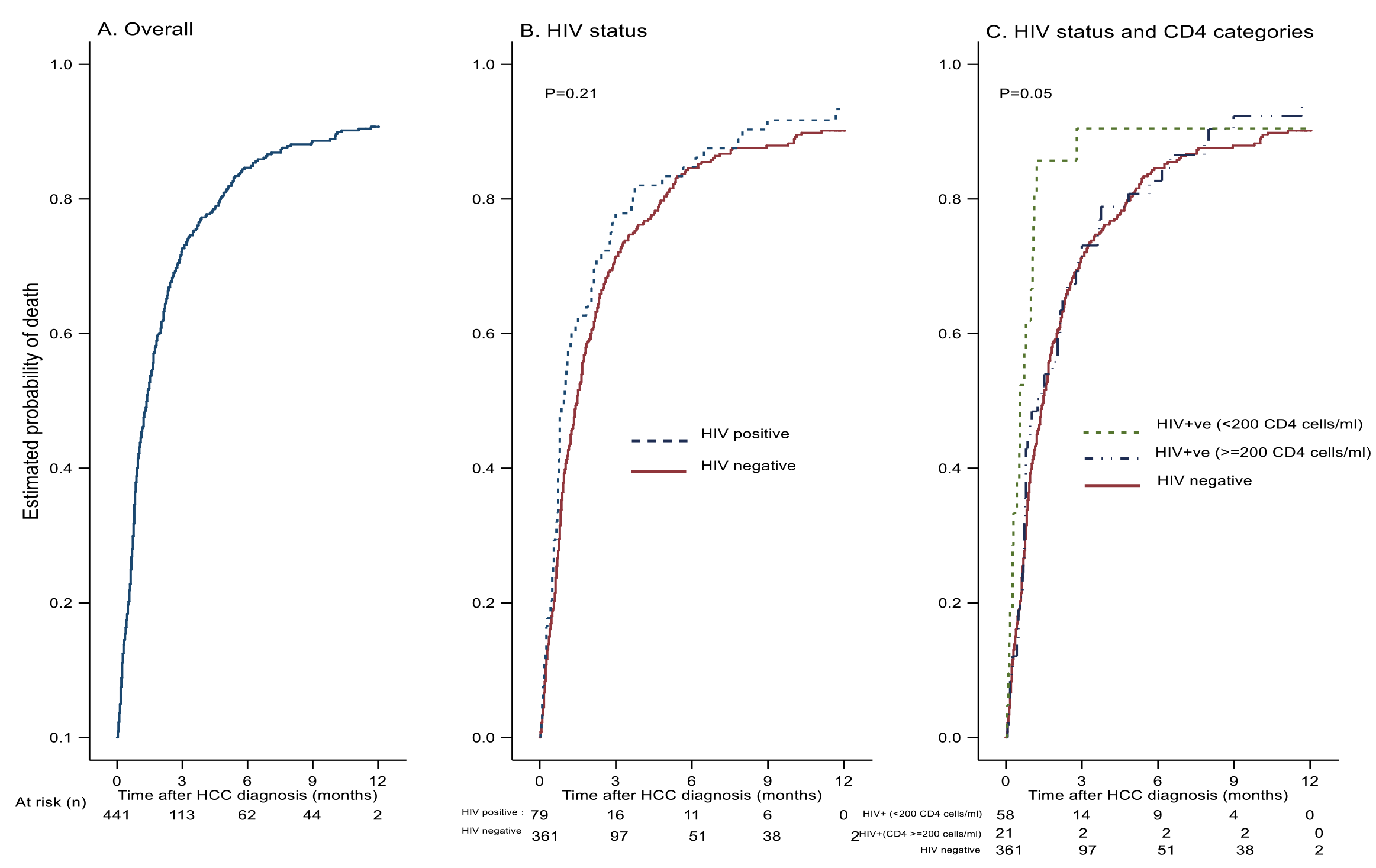
Survival after hepatocullar carcinoma diagnosis, overall, by HIV status, and by HIV and CD4 cell category. Figure 2 shows Kaplan-Meier curves depicting probability of death within 12 months after HCC diagnosis, (A) overall, (B) by HIV-infection status (p=0.21), (C) by HIV-infection status and CD4 cell categories (HIV-uninfected versus HIV-infected with ≥200 CD4 cells/ml versus HIV-infected with <200 CD4 cells/ml (p=0.05), and comparison of HIV-infected group with <200 CD4 cells/ml versus others (HIV-infected with ≥200 CD4 cells/ml or HIV-uninfected), (p=0.02). ^a^ p-value from non-parametric Log-rank test, a global test to compare if the survival between two or three independent groups, B and C respectively, are identical (overlapping) or not.

The crude hazard ratio (HR) and corresponding 95% confidence intervals (CI) of death after HCC diagnosis associated with HIV infection was 1.20 (0.93-1.56), p=0.16. In multivariable analysis adjusting for age, sex, education, alcohol and smoking history, HBsAg, anti-HCV and urine schistosomiasis CCA, HIV infection was associated with increased mortality risk although this difference did not achieve statistical significance, aHR (95% C) = 1.30 (0.97-1.74), p=0.081. Compared with HIV-uninfected counterparts, the higher mortality risk observed among HIV-infected individuals was significant among those with more severe HIV-disease (less than 200 CD4 cells per cubic milliliter), aHR (95% C) = 2.12 (1.23-3.53), p=0.004, but not among individuals with 200 or more CD4 cells per cubic milliliter, aHR (95% C) = 1.15 (0.82-1.60), p=0.417. In a similar trend of increased mortality risk associated with more advanced HIV disease albeit not statistically significant, compared to non-HIV HCC cases, higher mortality risk was observed among PLWH with detectable HIV viremia [aHR (95% C) = 1.47 (0.94-2.30) with VL ≥ 200 copies/ml”, p=0.095] than observed among those with current viral suppression [aHR (95% C) = 1.23 (0.87-1.73), p=0.243].

## Discussion

This investigation reveals the exceptionally high mortality associated with the diagnosis of HCC in sub-Saharan Africa. While there were no major differences detected in how HCC presented in PLWH, mortality was exceptionally fast in those with advanced HIV. Collectively, these data contribute to the urgency to identify and deploy new methods to prevent, detect and cure HCC worldwide.

HCC in sub-Saharan Africa, and particularly in Uganda, is one of the most common causes of cancer and of cancer-attributable death. Previously, we leveraged Kampala Cancer Registry data to identify an increasing trend in HCC incidence rates in Uganda which predated widespread availability of cART.^20^ Our suspicion, similar to what has been observed in more developed settings, is that HCC incidence rates are likely to continue to increase as PLWH survive into older ages in a setting with substantive chronic HBV infection and aflatoxin exposure. While this study does not calculate population-level HCC incidence rates, we do note that almost one in five HCC cases were HIV-infected, which is among the highest HIV prevalence reported of any prior non-targeted HCC study^21-23^.

Survival in patients with untreated advanced HCC is poor, although varying 1-year survival rates and prognostic factors have been reported depending on demographic and clinical characteristics, different timing of referral and diagnostic criteria, underlying cause of liver disease, severity of the underlying cirrhosis, and tumor burden.^24-26^ The short median survival after HCC diagnosis observed in this study is consistent with an Uganda study from three decades ago which characterized similar survival among untreated HCC patients^15^. Studies looking at survival in which participants received treatment for HCC disease have reported a relatively longer median survival beyond 10 months.^26-28^ Unfortunately, a recent review from sub-Saharan Africa indicates that only 3% of 1315 HCC cases from sub-Saharan Africa underwent any HCC specific therapy.^29^ Today, HCC in Africa represents an advance diagnosis with minimal treatment options with limited advancement over decades.

Despite the limited survival time, we observed an HIV effect towards more rapid mortality. This effect was generally observed by HIV status, but was accentuated when evaluating markers of more advanced HIV disease, notably CD4 count less than 200 cells/ml. Theoretically, PLWH engaged in a well-developed, HIV care delivery system in Uganda and achieving viral suppression should potentially be more readily identified at earlier HCC stages. Our data do not support this hypothesis however. Almost 80% of HCC cases in our study had multifocal liver lesions at diagnosis, irrespective of HIV status. The clinical signs, symptoms and laboratory studies of HCC cases also were very similar by HIV status. Despite this, PLWH with more advanced immunosuppression had to 30% more rapid time to death. We are unable to establish the underlying mechanism for advanced HIV disease resulting in earlier mortality, and advanced liver cirrhosis itself may contribute to changes in CD4 counts ^30^.

To our knowledge, this is the first African study to report the impact of HIV infection on survival after HCC diagnosis. Data from other regions are mixed; a study by Yopp et al from a single European institution reported that HIV infection was not associated with HCC survival.^31^. In contrast, other European studies^32,33^ showed a worse survival outcome among HIV-infected HCC patients; these studies were relatively small retrospective studies from tertiary care academic centers with possible biases of patient selection and treatment options. Despite several analyses of large-scale HIV and cancer datasets from the US that excluded HCC, ^34^ inferences from national data did find that PLWH with HCC presented at more advanced stages and had worse survival compared to non-HIV HCC even after accounting for a variety of health associated factors. In what purports to be among the largest international studies of the HIV effect on HCC survival (N=132 HIV associated HCC cases) from four continents (excluding sub-Saharan Africa), the authors estimated around the 25% increased mortality risk associated with HIV, despite appropriate ART and viral suppression.^35^

For 15% of HIV-associated HCC cases, the diagnosis of HCC prompted the initial identification of HIV infection. The small proportion of HCC cases who were unaware of their HIV status prior to study enrolment is consistent with data that has shown that Uganda has high rates of HIV testing and treatment.^36,37^ Around 77% of HIV-infected cases on cART had undetectable HIV viral loads which is similar to findings by Byonabye et al^38^ that have shown that PLWH in Uganda in care achieve high levels of viral suppression. Notably, these higher levels of viral suppression occurred despite only 57% of persons with advanced HCC disease reporting optimal ART adherence, emphasizing the potency of current ART regimens despite less-than-perfect adherence.

This study had both strengths and limitations. Many prior investigations of HCC in sub-Saharan Africa have been limited by the relatively small size and the reliance on convenience sampling or compilation of sporadic cases without standardized assessment of symptoms, clinical and laboratory findings or ultrasonographic features. Leveraging our standardized data collection procedures, we report uniformly-collected interview, clinical, laboratory, and ultrasonographic data among a large number of highly-characterized HCC cases systematically enrolled in a hospital-based study in Kampala, Uganda. The study was large with complete registration of participants, and though it was hospital-based, it was within a referral hospital with patients coming from several regions around the country with limited options for diagnosis and treatment for HCC. Despite the relatively large size for an HCC study from Africa, the numbers of cases were limited when evaluating stratified analyses by CD4 count or viral suppression status. Only a few of the patients with suspected HCC had a confirmatory histopathologic diagnosis and the possibility that other cancers such as cholangio-carcinoma or unspecified types being diagnosed as HCC is plausible. We limited misclassification of liver metastases as HCC by noting the differing ultrasonographic patterns and use of AFP, generally not elevated with metastases. Further, HCC accounts for approximately 70-90% of primary liver cancers^20,39,40^ in Uganda and in sub-Saharan Africa.

Although HCC cases with earlier stage symptoms can be identified, it remains unclear if more aggressive diagnostic approaches can identify earlier-stage HCC and improve survival. It is also important to start surveillance for HCC in high-risk patients including those with HIV infection in endemic sub-Saharan countries to ensure early diagnosis and thereafter linkage to localized curative treatments in order to modify the statistics caused by this lethal disease.

## Conclusions

Prognosis for HCC in sub-Saharan Africa is dismal and has largely remained the same over the last few decades^15,41^ Similar to the nihilism confronting HIV care delivery in sub-Saharan Africa prior to the widespread scale up of ART programs, substantial skepticism exists on whether HCC can be identified in earlier stages and curative treatments be provided to impact this current situation. However, global efforts have identified viral hepatitis elimination as a primary goal and increasingly, locally curative therapies and tumor resection (without transplantation) for earlier stage disease may become increasingly available. Within the past two years, the first liver resections for HCC have been performed in Uganda.

Advanced HIV disease appears to accelerate mortality among HCC patients with this already highly lethal disease. However, the existing HIV care infrastructure in sub-Saharan Africa where patients are engaged over long periods of time, advanced diagnostic testing is available, and antivirals can be successfully delivered provides a framework on which to leverage HCC surveillance programs. Leveraging this human and infrastructure capacity, resources should be invested to develop and evaluate the integration of active HCC surveillance programs within HIV care delivery systems, targeting both PLWH and high-risk, HIV uninfected persons, to identify patients with limited HCC amenable to locally available curative therapies.

## Data Availability

All data produced in the present study are available upon reasonable request to the authors.

## Declarations

### Ethics approval and consent to participate

This study was approved by the Institutional Review Boards of Makerere University School of Medicine and Johns Hopkins University. Written informed consent was obtained from all patients before participation in the study.

### Availability of data and materials

The datasets used and/or analysed during the current study are available from the corresponding author on reasonable request.

## Authors’ contributions

SN conceived the presented research idea and took the lead in writing the draft manuscript. AJ contributed to creating the final study dataset, reconstruction of study variables and took the lead in data analysis. GK, AJ and PO provided overall supervision and guidance of statistical analysis and contributed to revisions of the manuscript. GK,PO and DT conceptualized the study on which this manuscript is based, were the lead investigators and contributed to revisions of the manuscript.. All authors have read and approved the final manuscript.

## Acknowledgements

We thank the study participants and their caregiving families who agreed to participate in this study, and the study teams in Kampala and Gulu who have shown great dedication and resilience over the years. We wish to acknowledge the support from the Infectious Diseases Institute in Kampala, Uganda Cancer Institute, Mulago National Referral Hospital and St. Mary’s Hospital, Lacor, Uganda.

We would also like to thank Drs. Julie Nabweteme, Eve-Marie Benson and Fred Okuku for their contribution to this study.

